# Healthcare Performance of Leprosy Management in Peripheral Health Facilities of Dhanusa and Mahottari, Nepal

**DOI:** 10.1101/2022.12.28.22284024

**Authors:** Ram Kumar Mahato, Uttam Ghimire, Bijay Bajracharya, KC Binod, Deepak Bam, Deepak Ghimire, Uttam Raj Pyakurel, David TS Hayman, Basudev Pandey, Chuman Lal Das, Krishna Prasad Paudel

**Affiliations:** Epidemiology and Disease Control Division, Department of Health Services, Ministry of Health and Population, Kathmandu, Nepal; Epidemiology and Disease Control Division-Program Management Unit-SCI-GF, Nepal; Lalgadh Leprosy Hospital & Service center, Nepal Leprosy Trust, Lalgadh, Nepal; Seti Provincial Hospital, Dhangadhi, Nepal; Nepal Leprosy Fellowship, Jhapa/, Nepal; Molecular Epidemiology and Public Health Laboratory, Infectious Disease Research Centre, Hopkirk Research Institute, Massey University, Palmerston North, New Zealand; DEJIMA Infectious Disease Research Alliance, Nagasaki University, 1-12-4, Sakamoto, Nagasaki, Japan

**Keywords:** Leprosy Focal Person, Knowledge and skill scores, Documentation score, Case management scores

## Abstract

**Background:** The global elimination of leprosy transmission by 2030 is a World Health Organization (WHO) target. Nepal’s leprosy elimination program depends on early case diagnosis and the performance of health workers and facilities. The knowledge and skills of paramedical staff (Leprosy Focal Person, LFP) and case documentation and management by health facilities is therefore key to the performance of health care services.

**Methods:** The performance of health workers and facilities was evaluated through a combined cross sectional and retrospective study approach of 31 health facilities and their LFPs in Dhanusa and Mahottari Districts in Madhesh Province, Nepal. An average of 6 patients (paucibacillary, PB, or multibacillary, MB) per health facility registered within the 2018/2019 fiscal year were also enrolled in the study. LFP knowledge (e.g., of the three cardinal signs) and skills (e.g., nerve palpation) and facility processes (e.g., record keeping) were scored (e.g., 0, 1) and then rescaled to a proportion, where 1 is perfect. Internal benchmarking was used to guide performance management.

**Results:** Overall LFP knowledge and skill scores ranged from 0.16 to 0.63 (median 0.53, 95% confidence interval (CI), 0.46-0.6). Case documentation scores ranged from 0.15 to 0.87 (median 0.37, 95% CI 0.36-0.38), case management scores from 0.38 to 0.79 (median 0.54, 95% CI 0.53-0.55) and the overall healthcare scores ranged from 0.36-0.62 (median 0.48, 95% CI 0.47-0.49). Leprosy-related training was significantly related to the knowledge and skill of the health workers. All identified cases (n =187) adhered to the complete treatment and release after treatment (RFT) scheme, out of which 84.5% were satisfied with the service they were provided. Leprosy disability and Ear Hand and Feet (EHF) scores were not significantly reduced in treated patients during the study period, but counseling by LFPs significantly improved cases’ positive belief and practices regarding self-care.

**Conclusion:** Overall leprosy care performance was low (43%) and can be improved by evidenced-based training, onsite coaching, monitoring, and supervision to facilitate leprosy transmission elimination. The results highlight many of the challenges facing leprosy elimination programs.

## Introduction

Leprosy is an infectious, pathogenic disease of skin and peripheral nerves, caused by *Mycobacterium lepra* (1). The consequences of leprosy include physical disability and social stigma (2). The global prevalence and grade 2 disability of leprosy was recorded as 22.9 and 1.4 per million population in 2019 (3). In the same period, Nepal recorded 69 cases per million population, of which 5.45% (3.8 per million population) are grade 2 disabilities, despite the elimination of leprosy as national public health problem by 2009 (4,5). Nepal was one of the 16 counties that reported more than 1000 cases at the end of 2019 (6).

Leprosy diagnosis is based on the presence of at least one of three 61 cardinal signs: definite loss of sensation in a pale or reddish skin patch, a thickened or enlarged peripheral nerve with loss of sensation and/or weakness of the muscles supplied by that nerve, and the presence of acid-fast bacilli in a slit-skin smear (7). Multidrug therapy (MDT) is a key strategy to cure and prevent the transmission of leprosy.

The World Health Organization (WHO) set a target of leprosy transmission elimination by 2030 and cases reduced by 44% between 2005 and 2019 (6). Nepal, in line with the WHO’s roadmap to ‘zero leprosy’(8), has also set a target to achieve zero new autochthonous cases by 2030 (9,10). The elimination rate and early diagnosis of leprosy is associated with case management skills and attitudes of health service providers (11,12). The case management skills of peripheral health workers could be developed and improved by providing them with high quality training (13).

Multidrug therapy (MDT) is one of WHO’s effective strategies to prevent the transmission, cure leprosy infection, and limit and reverse patients’ disability status (14,15). Leprosy elimination programs therefore need high quality documentation to track the progress of cases and the programme (16,17). The Nepali National Leprosy Control Programme sets recording standards and provides the reporting tools in every health facility in a municipality. In order to record a case of leprosy, a health worker completes a Health Service Card (HMIS 1.2), Referral Form (HMIS 1.4), Laboratory Forms and Register (HMIS 5.1/5.2), Leprosy Diagnosis and Treatment Card (HMIS 5.4), Leprosy Treatment Register (HMIS 5.5), Defaulter Form (HMIS 1.5) and Reporting Form (HMIS 9.3)(18).

Finally, patient self-care is one of the main interventions to improve patient disability over time (19). Client satisfaction is a key performance indicator which improves self-care, drug adherence and recovery (20). Much of the information regarding care provided to leprosy cases is provided by paramedical staff LFP in Nepal.

This operational research was carried out in the Nepali context in Dhanusa and Mahottari districts, Nepal, to assess the knowledge and skills of LFPs and health facility overall performances. Metrics assessed include the documentation in health facilities, case diagnosis and management status, improvement in disability post-MDT, self-care practices in disabled people and client satisfaction, which reflects overall leprosy elimination performance. Gaps pertaining to knowledge, skill, documentation, and case management were identified and necessary interventions like strategic basic or refresher training, regular monitoring and supervision and onsite coaching were recommended.

## Material and methods

### Subjects and type of study

#### Location

This study was performed in Dhanusa and Mahottari districts in Madhesh Province, Nepal. Madhesh Province was chosen because it represents 40% of the notified leprosy cases in Nepal from the seven provinces, with Lumbini Province the next highest with 18% of cases.

#### Study design

A cross-sectional study to assess the knowledge and skills of LFPs was undertaken in 31 health facilities. A retrospective study of documentation and case management performance was undertaken to assess the overall performance of the respective health facilities. Thirty-one health municipalities were selected (17 from Dhanusha and 14 from Mahottari) based on convenience sampling, with just one municipality not sampled from each district (total health municipalities are 18 for Dhanusha and 15 for Mahottari); one health facility was then chosen randomly from each municipality. An LFP from each selected peripheral health facility was interviewed and assessed for their knowledge and skills pertaining to leprosy case management.

On average six patients (paucibacillary, PB, or multibacillary, MB) per health facility registered within the fiscal year 2018-19 and who had completed the treatment by the end of fiscal year 2019-20, were enrolled in the study. These patients were invited to enroll in the study by LFPs of respective health facilities. MB patients registered within the fiscal year 2018/2019 who had completed MB-MDT (12 blister packs) within 18 months, i.e., by the end of fiscal year 2019-20, were included. Similarly, PB patients registered within the fiscal year 2018-2019 and completed PB-MDT (6 blister packs) within 9 months before and by the fiscal year 2019/20 were included. In total, 187 patients were selected to assess the case management performance of the health facilities based on a sample size estimated to detect and improved performance (see below) among health facilities of 50%, with a precision of 1/7^th^ of 50% (± ∼7%) with 95% confidence.

#### Leprosy Focal Person skills and knowledge assessment

LFP knowledge of the 3 cardinal signs and capability to differentiate leprosy and non-leprosy patches was assessed through interviews by giving 12 patches (8 leprosy patches and 4 non-leprosy patches) from the leprosy atlas to all LFPs to identify correctly. An LFP was given 0.25 for correctly identifying up to 3 patches, 0.5 for 4-6 patches, 0.75 for 7-9 patches and 1 for 10-12 patches. The capability to palpate peripheral nerves (4 peripheral nerves: radial, ulnar, tibial, perineal nerves) was assessed by observation by a leprosy expert. A combined score of knowledge and skills was then given by providing equal weight to all variables, each given 1 mark and rescaled converted again to between 0 and 1.

#### Health facility documentation performance assessment

For the documentation performance, information on the completion and accuracy status of HMIS forms 5.4, 5.5, and 9.3 for the 187 patients from 31 facilities of Dhanusa and Mahottari was collected. Regarding part 1 of HMIS 5.4, a score was given as 0 for no charting at all and unavailability of the forms (hereon, performance), 0.33 for less than 1/3^rd^ performance, 0.67 for 2/3^rd^ performance and 1 for correctly and completely done charting. From parts 2 to 8 of HMIS 5.4, HMIS 5.5, HMIS 9.3, the score was calculated after assessing how accurately filled the variables were against the total variables. Similarly, randomly selected 3-month reports for each patient enrolled in the study were examined and given an average score for the documentation performance for each health facilities. Total score documentation was averaged and rescaled to between 0 and 1.

#### Case management performance assessment

For the case management performance of 187 patients from 31 health facilities, information on diagnosis accuracy, accuracy in classification of leprosy, treatment completion in time, number of doses taken as per the required treatment regime, disability and EHF score (pre– and post-treatment), number of follow-up visits, and counselling provided by the LFP with respect to treatment, self-care, attitude and practice of self-care, treatment outcome, reaction management by the LFP were collected. For all the patients, variables like accuracy in diagnosis, treatment completion in time, correct classification of disability, counselling provided by LFPs, practice of self-care by leprosy affected persons, reaction managed by LFP, completion of follow up visit, treatment outcome were treated as binomial variables; for the true value (i.e. correct classification, completed treatment, etc.), 1 mark was given and for the “not true” value, 0 was given. For post-treatment disability and EHF scores, 1 was given for improving, 0.5 for the same performance and 0 for worsening disability and EHF scores, with disability and EHF scores given separately. All post-treatment disability and EHF scores were summed and averaged and rescaled to between 0 and 1. Treated leprosy cases were asked if they were fully satisfied, satisfied or not satisfied with the services provided by the health system (health institution and health worker or LFP).

#### Performance summary

Finally, performance of health facilities was calculated by averaging the documentation scores and case management performance scores of respective health facilities and analyzing the data as above.

### Statistical analysis

Knowledge and skills of LFPs was analyzed by regression (see below) with predictors comprising their job title (Community Medical Assistant (one year training), Auxiliary Nurse Midwifery (1.5 years training course), Health Assistant (3-year academic course)), any training like basic leprosy training (BLT) and comprehensive leprosy training (CLT), and time since training taken. Similarly, pre– and post-disability and EHF score of leprosy treatment was analyzed.

Scores for the knowledge and skills of LFPs, documentation completion, and case management performance of health facilities were calculated in Microsoft® Excel. Means with 95% CI was calculated by using IBM SPSS 2022. Chi-squared (χ^2^) values, p-values and odds ratios with 95% CI were also calculated in SPSS and R version 4.2.0 to determine the association between knowledge and skill scores of LFPs and their leprosy training (BLT or CLT) and for the working designation compared with the knowledge and skill score of LFPs. Mean differences for disability and EHF scores related to leprosy were calculated pre– and post-leprosy treatment to know the impact of MDT in disability. Odds ratios and χ^2^ with 95% CIs were calculated for the association between status of regular counselling provided to leprosy cases and attitude and practices of selfcare. Case treatment satisfaction was calculated in terms of simple percentages of each satisfaction score for the service received from peripheral health facilities. Due to small sample sizes, univariate analyses are presented. However, beta regression (R^2^) was used to find predictors in variation in knowledge and skill scores of LFPs due to duration gaps after training and other factors. Specifically, knowledge and skills of LFPs were regressed against: training, qualification type and district and, separately, knowledge and skills of LFPs regressed against time from training. These regressions were done on raw data, not quantiles.

## Results

### Leprosy Focal Person backgrounds

The study assessed the knowledge and skills of 31 LFPs of selected peripheral health facilities of Dhanusa and Mahottari districts in Nepal. Among the selected 31 LFPs, 18 (58%, 39-75%) were community medical assistants (CMA), auxiliary nurse midwives (ANM) and auxiliary health workers (AHW) and 13 (42%, 25-61%) health assistants (HA). CMAs, ANMs and AHWs are qualified after a one and half year training course, whereas HAs are qualified after 3 years of an academic course. The study showed that more than half of the LFPs (16, 52%, 33-70%) have not taken any specific training. Very few LFPs (6, 19%, 7-37%) had graduate degrees of any discipline (Table S1).

### Leprosy Focal Person cardinal sign knowledge and palpation of peripheral nerve skills

Nine (29%, 14-48%) of LFPs knew all 3 cardinal signs, 8 (26%, 12-45%) knew two cardinal signs, 13 (42%, 25-61%) knew just one cardinal sign and 1 (3%, 0.1-17%) had no knowledge of cardinal signs of leprosy (Table S2). Further, 25 (81%, 63-93%) of the LFPs could not palpate any of the targeted 4 peripheral nerves, 4 (13%, 4-30%) of the LFPs could palpate one peripheral nerve, 1 (3%, 0.1-17%) LFPs could palpate 2 peripheral nerves, and just 1 (3%, 0.1-17%) could palpate all targeted peripheral nerves.

### Leprosy Focal Person knowledge and skill scores

Overall knowledge and skill scores ranged from 0.16 to 0.63 (median 0.53, 95% CI, 0.46-0.6; interquartile range 0.36-0.59) and the data were not normally distributed with some higher performances and then a spread below 0.5 (Figure 1).

**Figure 1.**
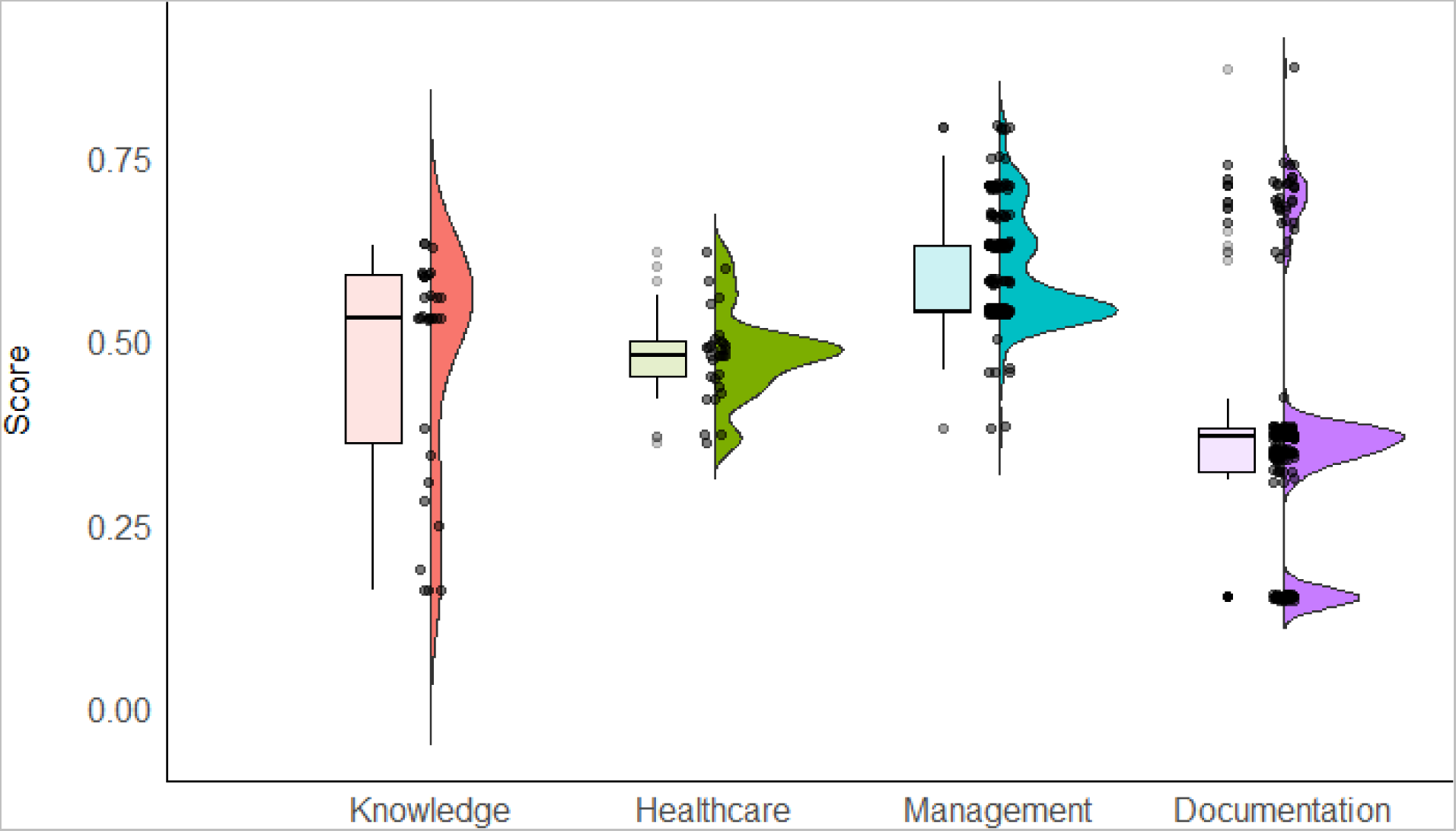
The leprosy focal person knowledge and skills and facility healthcare performance scores. Knowledge is the leprosy local person knowledge and skill scores; Healthcare the overall healthcare scores; Management the case management scores, and; documentation the case documentation scores. Box plots with outliers are shown with density plots with overlaid raw data to show the non-normal distributions.

### Predictors of Leprosy Focal Person knowledge and skill

Those who have taken any of the leprosy training courses (BLT or CLT) had significantly higher levels of knowledge and skills (OR = 8.1, 95% CI 1.4-66.3, p-value = 0.01). The level of knowledge and skills of LFPs was not significantly different with respect to their education level, i.e., a one-year training course (CMA or ANM or AHW) versus a 3-year academic course (HA) (p-value = 0.16). Neither 3-year HA qualifications (p-value = 0.82), nor receiving training (p-value = 0.46) significantly increased LFP knowledge and skills in multivariate regression, whereas Mahottari had significantly higher knowledge and skills than Dhanusa (β= 0.63, standard error (SE) = 0.21, p-value = 0.002) (Figure 2). Further, the time interval since the training was taken was not identified as predictor of knowledge and skills (Table S3; Figure S1, beta regression β=-0.007, SE=0.024, p-value = 0.78).

**Figure 2.**
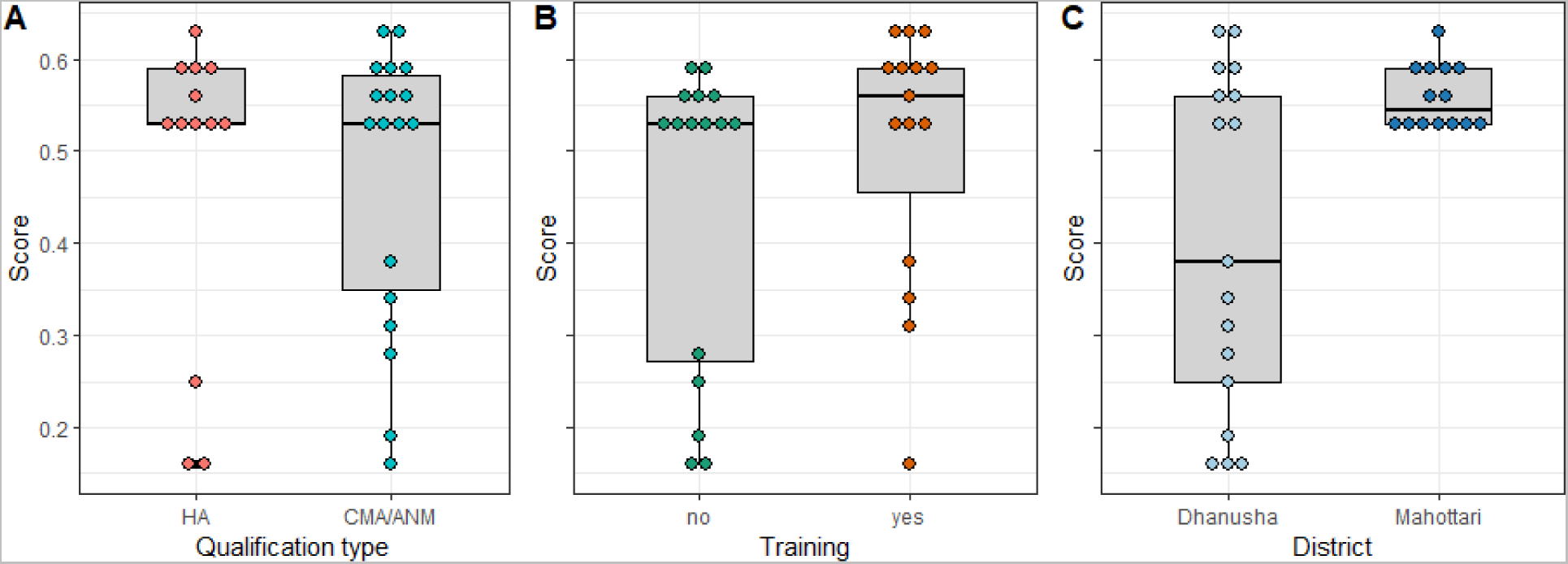
The distribution of leprosy focal person knowledge and skills scores by qualification type, training, and district. HA is health assistants (3-years of training); CMA community medical assistants; ANM auxiliary nurse midwives and auxiliary health workers (1-year of training).

### Health facility performance

All correctly diagnosed cases (N=187) completed treatment in time (N=184) and were released after the treatment (RFT, N=184), as per treatment guidelines. Three cases were under treatment, so they were excluded from the study when appropriate below, specifically the treatment analyses, and included in general analyses (i.e., case documentation and management analyses).

#### Case documentation

Case documentation scores ranged from 0.15 to 0.87 (Figure 1). The median score was 0.37 (95% CI 0.36-0.38; IQR 0.32-0.38) and the data were not normally distributed with 75 (40%, n=187) considered outliers and many scores much higher or lower than this mean. (Figure 1).

#### Case management scores

Case management scores ranged from 0.38 to 0.79 (Figure 1). The median score was 0.54 (95% CI 0.53-0.55; IQR 0.54-0.63) and again the data were not normally distributed with 5 outliers.

### Overall healthcare scores

Overall healthcare scores ranged from 0.36-0.62 (Figure 1). The median score was 0.48 (95% CI 0.47-0.49; IQR 0.45-0.5) and again the data were not normally distributed with 5 outliers.

### Place of leprosy case diagnosis

More than 3/4 (145, 78%, 71-83%) of the cases were diagnosed in Lalgadh Leprosy Hospital, in Dhanusa (Table S4). Less than one fifth (32, 17%, 12-23) cases were diagnosed in government peripheral health facilities. Just nine (5%, 2-9%) of the leprosy cases were diagnosed through active case detection, i.e., a contact survey by the Leprosy Post-Exposure Prophylaxis (LPEP) service. One case (<1% cases, <0.1-3%) was diagnosed in India, to which Dhanusa and Mahottari are bordering districts.

### Pre– and post-disability status of MDT treated patients

No statistical improvement in the disability status or EHF score at the time of study in MDT treated patients compared to the disability and EHF score recorded at the time of registration or before the treatment was measurable (95% CI –6 to 12) (Table 1; Figure S2).

**Table 1.** Disability and Ear Hand and Feet (EHF) score in Multidrug therapy treated patients.

| Score at the time of treatment |  | Score at the time of study |  | Statistics |
| --- | --- | --- | --- | --- |
| Disability (grade 1 or 2) |  |  |  |  |
| None | Present | None | Present |  |
| 143 (76%, 70-82%) | 44 (23%, 18-31%) | 148 (79%, 73-85%) | 39 (21%, 15-27%) | $\chi^2 = 0.25$, df = 1, p-value = 0.62; 95% CI change: -6 to 12% |
| EHF |  |  |  |  |
| 0 | >1 | 0 | >1 |  |
| 143 (76%, 70- | 44 (23%, 18- | 149 (80%, 73- | 38 (20%, 15- | $\chi^2 = 0.39$, df = 1, p-value = |
| 82%) | 31%) | 85%) | 27%) | 0.53; 95% CI change: -6 to<br>12% |

### The role of counselling for self-care after RFT in improving disability

Overall, self-care did not significantly improve disability (β=0.25, SE=0.26, p-value = 0.34) or EHF (β=0.45, SE = 0.35, p-value =0.21) scores (Figure S3). However, patients who have been provided counselling for self-care know that self-care can improve disabilities significantly better than those who have not been provided with the counseling for self-care (OR 7.9, 95% CI, 1.3-89.1, p-value = 0.01). Moreover, those patients who have not been provided with counselling practice self-care significantly less than those who have been provided with counselling (OR = 46 (95% CI, 2.5-867%), p-value <0.001) (Table S5).

### Lepra reaction status ever experienced in leprosy treated patients

Most of the patients (172, 92%, 87-95%) had no lepra reaction ever, whereas just 5 (2.7%, 1-6%) had had a type 1 reaction and 10 (5.4%, 3-10%) a type 2 reaction. Of the 15 people with a positive lepra reaction, just one (6.7%, 0.2-32%) was a case being managed by an LFP.

### Client satisfaction status in leprosy treated patients

Only 20 (11%, 7-16%) of leprosy treated patients was fully satisfied with the health services, yet more than 4/5 (158, 85%, 78-89%) of the patients were satisfied with the services. Similarly, only a few (9, 5%, 2-8%) were not satisfied with the services.

## Discussion

The Nepal country roadmap to zero leprosy recommends high levels of expertise to facilitate the early detection and reduction in transmission of leprosy (9,21). Contrary to that, the median score of knowledge and skills of LFPs in Dhanusa and Mahottari was just 0.53. The low documentation score of 0.37 in the study districts is a challenging issue, because high quality documentation is required to assess the progress and validate the elimination of leprosy (22). The exact situation of leprosy elimination in Dhanusa and Mahottari was therefore likely uncertain, as the median case management and median overall performance was just 0.54 and 0.48 respectively.

Fifty percent of LFPs had knowledge and skills less than 53%, which is low and is in contrast Mohite *et al*’s study in Bangladesh, where 88% of health workers had good knowledge of leprosy (20). The highest scores were just 63%. Similarly, most peripheral health facilities had poor performance scores posing a serious concern for elimination of transmission of leprosy by the year 2030.

Ninety six percent of the LFPs knew at least 1 cardinal sign, a more satisfactory result compared to a study in Bangladesh (72%) (23). However, only 19% of LFPs could palpate at least one peripheral nerve, revealing gaps in leprosy diagnosis in Nepal which need addressing. Only 19% (6 out 31) health facilities received a 50% aggregate performance regarding leprosy management, crucial for achieving the goal of elimination of transmission by 2030. The finding that >80% of the LFPs could not palpate any peripheral nerves is comparable to the study conducted by Roy *et al* that showed 83% health workers in parts of India were poorly skilled (13).

Those workers who have taken any of the leprosy training (BLT or CLT) had significantly higher levels of knowledge and skill (OR = 8.1, 95% CI, 1.4-66.3, p-value = 0.01), highlighting the importance of training since leprosy is either not included or not prioritized in the academic course of paramedical staff. Other variables including education level and duration of experience were not significant markers of increased knowledge and skills. However, in multivariate regression of the raw scores, Mahottari had significantly higher knowledge and skills than Dhanusa (β= 0.63, standard error (SE) = 0.21, p-value = 0.002; Figure 2), but not training. This needs to be explored further.

Less than 5% (4.8%) of cases were diagnosed through active case detection and the majority of cases (78%) were diagnosed through the Lalgadh Leprosy Hospital, which drew the governments’ attention to hidden cases in the community. The cases are diagnosed at the specialty hospitals only after the leprosy cases reach the advanced stage. This means the proportion of the cases diagnosed at the peripheral health facilities should be greater, as people in the early stages of leprosy visit these facilities. In this study, few leprosy cases were diagnosed in the peripheral health facilities. So, early cases could be hidden in community due to the inefficient performance of health worker, unavailability of diagnosis and/or the stigma associated with the disease.

Both disability score and EHF score were not significantly improved after full treatment and compliance with MB MDT treatment, which was surprising as comparable studies conducted by Kumar *et al* show as the disability prevalence in non-compliant cases was significantly greater than fully treated patients (15). This may, however, be due to the very small sample size and time frame and requires further investigation. That all cases have completed treatment in time and were released after treatment indicates a well performing health system for those recruited. Moreover, counselling of patients by a LFP had significantly changed the belief of the patient that self-care can improve disability (OR = 7.9, 95% CI, 1.3-89.1, p-value = 0.01). The result was comparable with the studies conducted by Lusli *et al* and Devkota *et al* (24,25), where counselling has a significant role in changing the belief and attitude towards health behaviors. Similarly, counseling significantly improved self-care practice (OR = 46, 95% CI, 2.5-867%, p-value = <0.001).

Among the cases monitored, only 8% developed lepra reactions, of which 2.7% (5 patients) and 5.4% (10 patients) were type 1 and type 2 reactions respectively. Other studies found 17.9% of leprosy cases developed type 1 reactions and between 1.2% to 15.4% of the leprosy cases developed type 2 reactions (26,27). These are similar trends but likely limited by sample sizes.

Regarding the satisfaction from serviced provided from health facilities, 95.2% leprosy cases were satisfied with the services, which is comparable to a study in Brazil, where between 92-100% of patients were satisfied at some level (28).

Weaknesses of our study are the convenience sampling of the districts, though this was deliberate due to disease burden, and patient enrolment by the LFPs of respective health facilities. This might have biased the sample and future prospective and randomized control studies might improve this. Moreover, the sample size is low due to low case numbers, potentially preventing more detailed analyses. For example, we used univariate count data analyses where continuous data and multivariate analyses would be beneficial, but we had too many variables for the small sample size. Similarly, we had a large, zero-inflated sample, with many patients having zero disability or EHF score. Zero-inflated beta regression tools exist and could be used, but failed to fit to our data, even using a Bayesian framework (not shown), likely due to too many zeros and prior distribution specification sensitivity. These factors limit some of the conclusions that can be made highlights the need for more comprehensive study.

## Conclusion

The overall performance of health care services for leprosy patients was 0.48 (95% CI, 0.47-0.49), in which median knowledge and skill scores of LFPs, documentation scores and case management score of health facilities were 0.53 (0.46-0.60), 0.37 (0.36-0.38) and 0.54 (0.53-0.55) respectively. These low scores likely hinder the elimination of leprosy transmission targets, despite all cases utilizing treatment and being released after treatment and 95.2% of cases being satisfied with the service they were provided. Leprosy-related training was significantly related to knowledge and skill of the health workers, which provides an evidence-base for further training. Disability and EHF scores were not significant reduced in treated patients in the study period, but counseling by LFP has significantly improved positive belief and practices regarding self-care, again offering evidence that training can help cases and potentially will contribute to reducing onward transmission. As the overall performance leprosy care service was 48%, it should be improved by intense evidenced-based training, onsite coaching, monitoring and supervision in order to facilitate elimination of transmission of leprosy by the year of 2030. The compliance should be maintained regularly, and this study provides a benchmark against which progress can be assessed.

## Acknowledgments

The authors would like to acknowledge, Dr Krishna Lama, Director of Lalgadh Leprosy Hospital, Mr. Bijay Kumar Jha, Director and Mr. Rajbir Yadav, focal people of the leprosy program from Madhesh Province, as well as chief and LFPs from the health office, Dhanusha and Mahottari, and section chiefs of the selected municipalities, as well as LFPs from all health facilities for their support and valuable inputs, and Drs Rosemary Barraclough and Jonathan Marshall, Massey University, for their useful feedback.

## Contributors

RKM conceived and designed the manuscript; RKM, UG, DG, DB, DKC managed and collected the data collection; RKM, DTSH analyzed the data; RKM, BB wrote the original draft, KPP, CLD, URP, DTSH, BDP reviewed the manuscript. All authors read, reviewed, and approved the final manuscript. DTSH is supported by the Percival Carmine Chair in Epidemiology and Public Health and by Royal Society Te Apūrangi RDF-MAU1701

## Data availability

The datasets generated and/or analyzed during the current study are not publicly available due to individual privacy of patients could be compromised but are available from the corresponding author on reasonable request.

## Ethics approval

The data were generated through monitoring, supervision and surveillance of health care performance as part of regular program activities by the Epidemiology and Disease Control Division (EDCD). While conducting the field surveillance, consent from the participants was taken verbally in the presence of local health authorities. Then all acquired field data were stored at the central database as per the national surveillance system. For this study, anonymized data were used and approved for analysis and publication by **Ethical Review Board of Nepal Health Research Council** (**approval number 349/2022 P and approval date: 21 July 2022**) as per the request from EDCD.

## Abbreviations

AHW: Auxiliary health worker
ANM: Auxiliary nurse midwives
BLT: Basic leprosy training
CI: Confidence Interval
CLT: Comprehensive leprosy training
CMA: Community medical assistant
EHF: Ear hand and feet
HMIS: Health Management Information System
LFP: Leprosy focal person
LPEP: Post-exposure prophylaxis
MB: Multibacillary
PB: Paucibacillary
RFT: Release after treatment
SPSS: Statistical package for the social sciences
WHO: World Health Organization

## References

1. Lastória JC, de Abreu MAMM. Leprosy: Review of the epidemiological, clinical, and etiopathogenic aspects – Part 1. An Bras Dermatol. 2014;89(2):205–18.

2. Joseph GA, Sundar Rao PSS. Impact of leprosy on the quality of life. Int J Lepr Other Mycobact Dis. 1999;67(4 SUPPL.):518.

3. World Health Organization (WHO). Leprosy (Hansen’s disease) [Internet]. 2021. p. 1–5. Available from: https://www.who.int/news-room/fact-sheets/detail/leprosy

4. Ministry of Health and Population (MoHP). Leprosy Control Programme (/ eng / program/ communicable [Internet]. 2015. Available from: https://www.mohp.gov.np/eng/program/communicable-disease/leprosy-control-program

5. Department of Health services(DoHS). Annual Report: Department of Health Services 2075/76 (2018/19) [Internet]. Vol. 76, Departement of Health Services, Ministry of Health and Population, Government of Nepal. 2019. Available from: https://publichealthupdate.com/department-of-health-services-dohs-annual-report-2075-76-2018-19/

6. Strategy GL. Global consultation of National Leprosy Programme managers, partners and affected persons on Global Leprosy Strategy. 2021.

7. Harrell GT. Epidemiology of leprosy. J Am Med Assoc. 1947;135(11):732.

8. WHO. Towards zero leprosy Global Leprosy (Hansen’s disease) Strategy 2021-2030. Word Heal Organ. 2021;1–30.

9. Department of Health services(DoHS). COUNTRY ROADMAP FOR ZERO LEPROSY. 2021.

10. National Planning Commission. National Planning Commission, 2015: Sustainable Development Goals, 2016-2030, National (Preliminary) Report. Government of Nepal, National Planning Commission, Kathmandu, Nepal. 2015. 39–52 p.

11. Pal R, Kar S, Ahmad S. Current knowledge attitudes, and practices of healthcare providers about leprosy in Assam, India. J Glob Infect Dis. 2010;2(3):212.

12. Ukpe I. A study of health workers’ knowledge and practices regarding leprosy care and control at primary care clinics in the Eerstehoek area of Gert Sibande district in Mpumalanga Province, South Africa. South African Fam Pract. 2006;48(5):16–16e.

13. Roy R, Shrivastava P, Mitra K, Mondal S. Assessment of Knowledge and Skills of Peripheral Health Workers for Screening and Detection of Early Nerve Damage in Leprosy□: A Cross-Sectional Assessment of Knowledge and Skills of Peripheral Health Workers for Screening and Detection of Early Nerve Dam. J Med Dent Sci. 2018;17(6):5–9.

14. World Health Organization. The final push strategy to eliminate leprosy as a public health problem. Quest answers [Internet]. 2003;73(3):279–81. Available from: http://www.who.int/lep/resources/Final_Push_QA.pdf

15. Kumar A, Girdhar A, Kumar Girdhar B. Risk of developing disability in pre and post-multidrug therapy treatment among multibacillary leprosy: Agra MB Cohort study. BMJ Open. 2012;2(2):1–7.

16. Department of Health services(DoHS). The Leprosy Post-Exposure Prophylaxis (LPEP) programme: Update and interim analysis. Lepr Rev [Internet]. 2018;89(2):102–16. Available from: https://www.researchgate.net/publication/326231998_The_Leprosy_Post-Exposure_Prophylaxis_LPEP_programme_Update_and_interim_analysis%0Ahttps://www.scopus.com/inward/record.uri?eid=2-s2.0-85052234154&partnerID=40&md5=47e0d137909d005f141a14e5d0a31263%0Ahttp:

17. Mori S, Barua S, Suzuki K, Yotsu RR, Ishii N. [Present leprosy situation in the world in 2012]. Nihon Hansenbyo Gakkai Zasshi. 2013;82(1–2):59–69.

18. Ministry of Health and Population (MoHP). Health Management Information System Guidelines, 2017,. Heal Manag Inf Syst Guidel Teku Kathmandu, Nepal [Internet]. 2017; Available from: https://dohs.gov.np/hmis-guideline-2017/

19. Eze CC, Ekeke N, Alphonsus C, Lehman L, Chukwu JN, Nwafor CC, et al. Effectiveness of self-care interventions for integrated morbidity management of skin neglected tropical diseases in Anambra State, Nigeria. 2021;1–15.

20. Mohite R V., Mohite VR, Durgawale PM. Patient satisfaction in national leprosy eradication programme. Bangladesh J Med Sci. 2013;12(3):305–9.

21. Subedi M, Engelbrektsson U-B. Factors Contributing to Delay in Diagnosis and Start of Treatment of Leprosy: Analysis of Help-seeking Narratives from a Community Study in Dang District. Dhaulagiri J Sociol Anthropol. 2018;12:11–7.

22. World Health Organization. Defining criteria to declare elimination of leprosy. Report of an informal consultation. 2020. 10–12 p.

23. Kabir H, Hossain S. Knowledge on leprosy and its management among primary healthcare providers in two districts of Bangladesh. BMC Health Serv Res. 2019;19(1):1–9.

24. Devkota R, Khan GM, Alam K, Sapkota B, Devkota D. Impacts of counseling on knowledge, attitude and practice of medication use during pregnancy. BMC Pregnancy Childbirth. 2017;17(1):1–7.

25. Lusli M, Peters R, van Brakel W, Zweekhorst M, Iancu S, Bunders J, et al. The Impact of a Rights-Based Counselling Intervention to Reduce Stigma in People Affected by Leprosy in Indonesia. PLoS Negl Trop Dis. 2016;10(12):1–25.

26. Voorend CGN, Post EB. A Systematic Review on the Epidemiological Data of Erythema Nodosum Leprosum, a Type 2 Leprosy Reaction. PLoS Negl Trop Dis. 2013;7(10).

27. Rosdiana B, Astari L, Astindari A, Prakoeswa CRS, Zulkarnain I, Damayanti D, et al. Risk factors of type 1 leprosy reaction in leprosy patients attending leprosy division of dermatology and venereology outpatient clinic of dr soetomo general hospital in 2017– 2019: A retrospective study. Open Access Maced J Med Sci. 2021;9:1359–63.

28. Ferreira IP oliann S, Buhrer-Sékula S, De Oliveira MR egin. F, Gonçalves H de S, Pontes MA rac. de A, Penna ML úci. F, et al. Patient profile and treatment satisfaction of Brazilian leprosy patients in a clinical trial of uniform six-month multidrug therapy (U-MDT/CT-BR). Lepr Rev. 2014;85(4):267–74.

