## Supplement for "Healthcare Performance of Leprosy Management in Peripheral Health Facilities of Dhanusa and Mahottari, Nepal"

**Supplementary information**

**Tables**

**Table S1. Background of Leprosy Focal Persons**

| **Background characteristics** | **Number and % (95% CI) of focal persons (N = 31)** |
| --- | --- |
| CMA or ANM (no academic course, but 1 year health training) | 18 (58%, 39-75%) |
| HA (completed 3 years academic training) | 13 (42%, 25-61%) |
| Leprosy specific training (either BLT or CLT) | 15 (48%, 30-67%) |
| Graduate degrees | 6 (19%, 7-37%) |
| Undergraduate degrees | 25 (81%, 63-93) |

**Table S2. Focal person’s knowledge on leprosy cardinal signs and skills on palpation of peripheral nerves**

| **Details** | **Numbers (%, 95% CI, N=31)** | **Details** | **Numbers (%, 95% CI, N=31)** |
| --- | --- | --- | --- |
| Persons who know 3 cardinal signs | 9 (29%, 14-48%) | Persons who palpate 4 peripheral nerves | 1 (3%, 0.1-17%) |
| Persons who know 2 cardinal signs | 8 (26%, 12-45%) | Persons who palpate 3 peripheral nerves | 0 (0%, 0-11%) |
| Persons who know only 1 cardinal sign | 13 (42%, 25-61%) | Persons who palpate 2 peripheral nerves | 1 (3%, 0.1-17%) |
| Persons who know no cardinal signs | 1 (3%, 0.1-17%) | Persons who palpate only 1 peripheral nerve | 4 (13%, 4-30%) |
|  |  | Persons who palpate no peripheral nerves | 25 (81%, 63-93%) |

**Table S3. Predictors of Leprosy Focal Person knowledge and skills**

| **Tested predictor** | **Level of knowledge and Skills**  **Statistics** | | **Statistics** |
| --- | --- | --- | --- |
|  | # Health workers above the median | # Health workers below the median |  |
| Training status |  |  |  |
| Training taken | 12 | 3 | OR = 8.1 (95% CI, 1.4-66.3), p-value = 0.01 |
| Training not taken | 5 | 11 |  |
| Job title |  |  |  |
| CMA, ANM or AHW | 12 | 6 | OR = 3.1 (95% CI, 0.6-18.3), p-value = 0.16 |
| HA | 5 | 8 |  |
| Time since training |  |  |  |
| ≤3 years | 3 | 6 | OR = 0.22 (95% CI, 0.01-2.45), p-value = 0.31 |
| >3 years | 5 | 2 |  |

**Table S4. Distribution of cases treated in Health facilities as per place for diagnosis**

| **Description** | **Number of cases (N=187) (%, 95% CI)** |
| --- | --- |
| Lalgadh Leprosy Hospital (specialist hospital) | 145 (78%, 71-83%) |
| Government local health facilities | 32 (17%, 12-23) |
| Contact survey, LPEP –Active case detection | 9 (5%, 2-9%) |
| India | 1 (0.5%, <0.1-3%) |

**Table S5. The perception and practice of self-care in leprosy disabled persons after RFT**

| **Counselling status** | **Patient status** | | **Statistics** |
| --- | --- | --- | --- |
|  | Believes self-care improves disability | Believes self-care does not improve disability |  |
| Provided | 23 | 2 | OR = 7.9 (95% CI, 1.3-89.1), p-value = 0.01 |
| Not provided | 11 | 8 |  |
|  | Practiced self-care | Did not practice self-care |  |
| Provided | 25 | 0 | OR* = 46 (95% CI, 2.5-867%), p-value = <0.001 |
| **Not provided** | **10** | **9** |  |

*Note: OR and 95% CIs were calculated using the Haldane correction due a zero (see Supplementary information equation), fisher.test in R was used to calculate the p-value.

**Equation**

**Equation S1** Odds ratio 95% confidence calculation with the Haldane correction:

$$e^{ln(OR)\pm1.96*\sqrt{1/(a+0.5)+1/(b+0.5)+1/(c+0.5)+1/(d+0.5)}}$$

**Figures**

**Figure S1** Leprosy focal persons knowledge and skills score and time of training

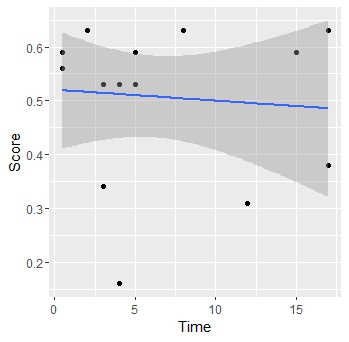

**Figure S2** Time and EHF (A) and disability scores (B).

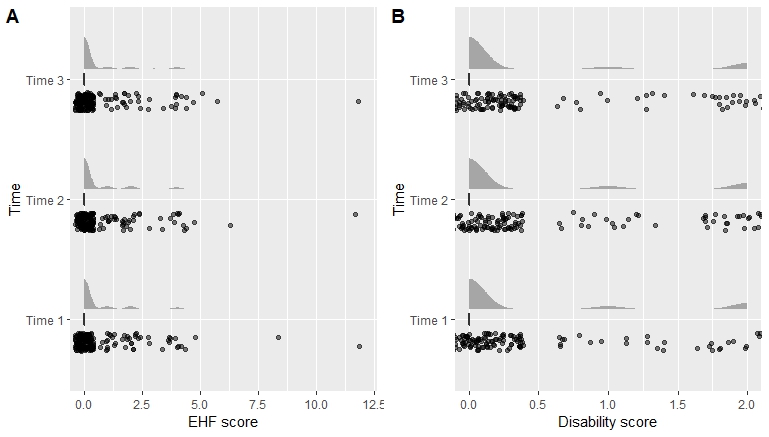

**Figure S3** Self-care and disability and ear, hand and feet score data

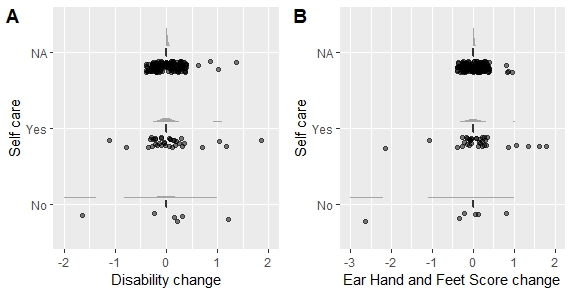
